# Identifying and Ranking Common COVID-19 Symptoms from Arabic Twitter

**DOI:** 10.1101/2020.06.10.20127225

**Authors:** Eisa Alanazi, Abdulaziz Alashaikh, Sarah Alqurashi, Aued Alanazi

**Affiliations:** Center of Innovation and Development in Artificial Intelligence, Umm Al-Qura University, Saudi Arabia; Department of Computer Engineering and Network, University of Jeddah, Saudi Arabia

**Author notes:** **Corresponding Author:** Eisa Alanazi, BSc MSc PhD, Center of Innovation and Development in Artificial Intelligence, Umm Al-Qura University, Makkah, 21421, Saudi Arabia.

**Keywords:** Social networks analysis, Twitter, data mining, covid-19, early symptoms, ranking, Arabic

## Abstract

**Background:** Massive amount of covid-19 related data is generated everyday by Twitter users. Self-reports of covid-19 symptoms on Twitter can reveal a great deal about the disease and its prevalence in the community. In particular, self-reports can be used as a valuable resource to learn more about the common symptoms and whether their order of appearance differs among different groups in the community. With sufficient available data, this has the potential of developing a covid-19 risk-assessment system that is tailored toward specific group of people.

**Objective:** The aim of this study is to identify the most common symptoms reported by covid-19 patients in the Arabic language and order the symptoms appearance based on the collected data.

**Methods:** We search the Arabic content of Twitter for personal reports of covid-19 symptoms from March 1^st^ to May 27^th^, 2020. We identify 463 Arabic users who tweeted testing positive for covid-19 and extract the symptoms they publicly associate with covid-19. Furthermore, we ask them directly through personal messages to opt in and rank the appearance of the first three symptoms they experienced right before (or after) diagnosed with covid-19. Finally, we track their Twitter timeline to identify additional symptoms that were mentioned within ±5 days from the day of tweeting having covid-19. In summary, a list of 270 covid-19 reports were collected and symptoms were (at least partially) ranked from early to late.

**Results:** The collected reports contained roughly 900 symptoms originated from 74% (n=201) male and 26% (n=69) female Twitter users. The majority (82%) of the tracked users were living in Saudi Arabia (46%) and Kuwait (36%). Furthermore, 13% (n=36) of the collected reports were asymptomatic. Out of the users with symptoms (n=234), 66% (n=180) provided a chronological order of appearance for at least three symptoms.

Fever 59% (n=139), Headache 43% (n=101), and Anosmia 39% (n=91) were found to be the top three symptoms mentioned by the reports. They count also for the top-3 common first symptoms in a way that 28% (n=65) said their covid journey started with a Fever, 15% (n=34) with a Headache and 12% (n=28) with Anosmia. Out of the Saudi symptomatic reported cases (n=110), the most common three symptoms were Fever 59% (n=65), Anosmia 42% (n=46), and Headache 38% (n=42).

**Conclusions:** This study demonstrates that Twitter is a valuable resource to analyze and identify COVID-19 early symptoms within the Arabic content of Twitter. It also suggests the possibility of developing a real-time covid-19 risk estimator based on the users’ tweets.

## Introduction

The ongoing vigorous COVID-19 outbreak has shown a great impact on human health and well-being and radically enforced a rigorous change in societies lifestyle undermining their prosperity. Along with this catastrophe, we have witnessed a great effort from diverse research communities to study this disease in all its aspects.

In recent years, social networks have become an unignorable source of information where users expose and share ideas, opinions, thoughts, and experiences on a multitude of topics. Several researches have utilized the abundance of information offered by social platforms to conduct non-clinical medical research. For example, Twitter has been the source for data for many health and medical studies; such as surveillance and monitoring of Flu and Cancer timeline and distribution across the USA using Twitter [1], analyzing the spread of influenza in the UAE based on geo-tagged Arabic Tweets [2], surveillance and monitoring of Influenza in the UAE based on Arabic and English tweets [3], identifying symptoms and disease in Saudi Arabia using Twitter [4], and most recently on analyzing COVID-19 symptoms on Twitter [5].and analyzing the chronological and geographical distribution of COVID-19 infected tweeters in the USA [6].

Twitter platform enables obtaining multiple features (such as age, sex, geo-location, … etc.) along with informative messages that using appropriate data mining and analysis techniques can potentially result in useful insights about a specific health condition [7]. Extracting common symptoms associated with a disease from publicly available data has the potential to control the spread of the disease and identify users at high risk. It also gives new insights that call for early intervention and control. For example, Figure 1 highlights a tweet (from Saudi Arabia) mentioning explicitly the loss of smell and taste as one distinctive symptom of covid-19. The national-wide self-testing questionnaire App was updated almost couple of weeks after the tweet to include the loss smell and taste as one potential sign for covid-19 [8]. Tracking covid-19 symptoms in real-time from public data on Twitter could have shorten the gap.

**Figure 1:**
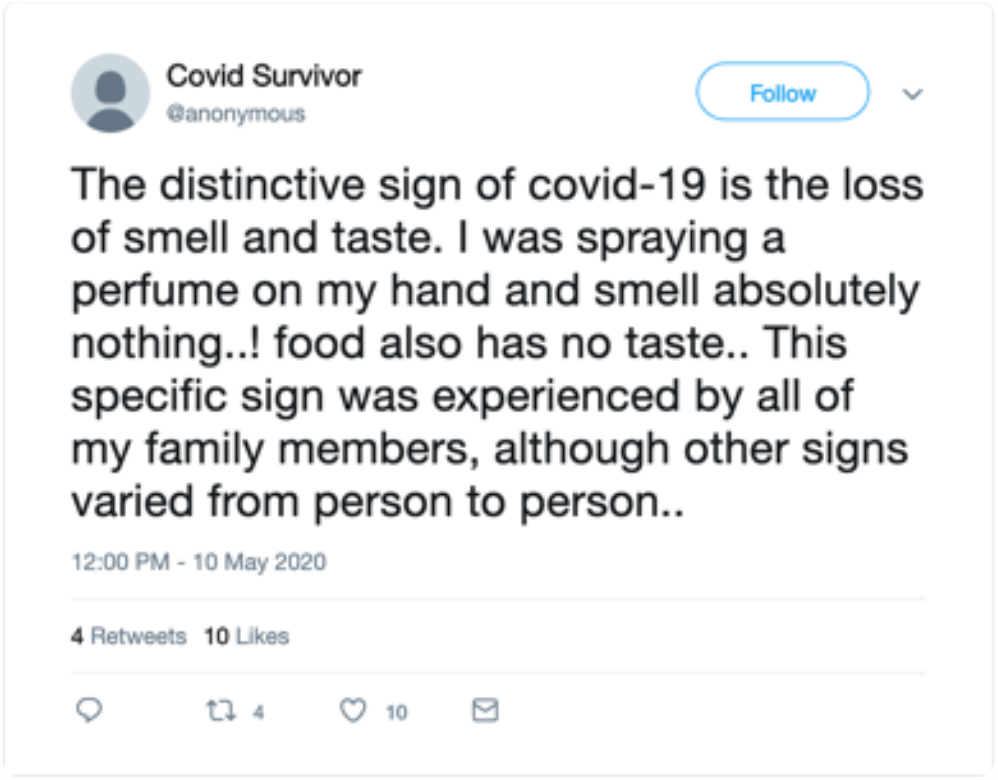
A covid-19 survivor tweets about how the loss of smell and taste was the only common sign among all of her family members.

**Figure 2:**
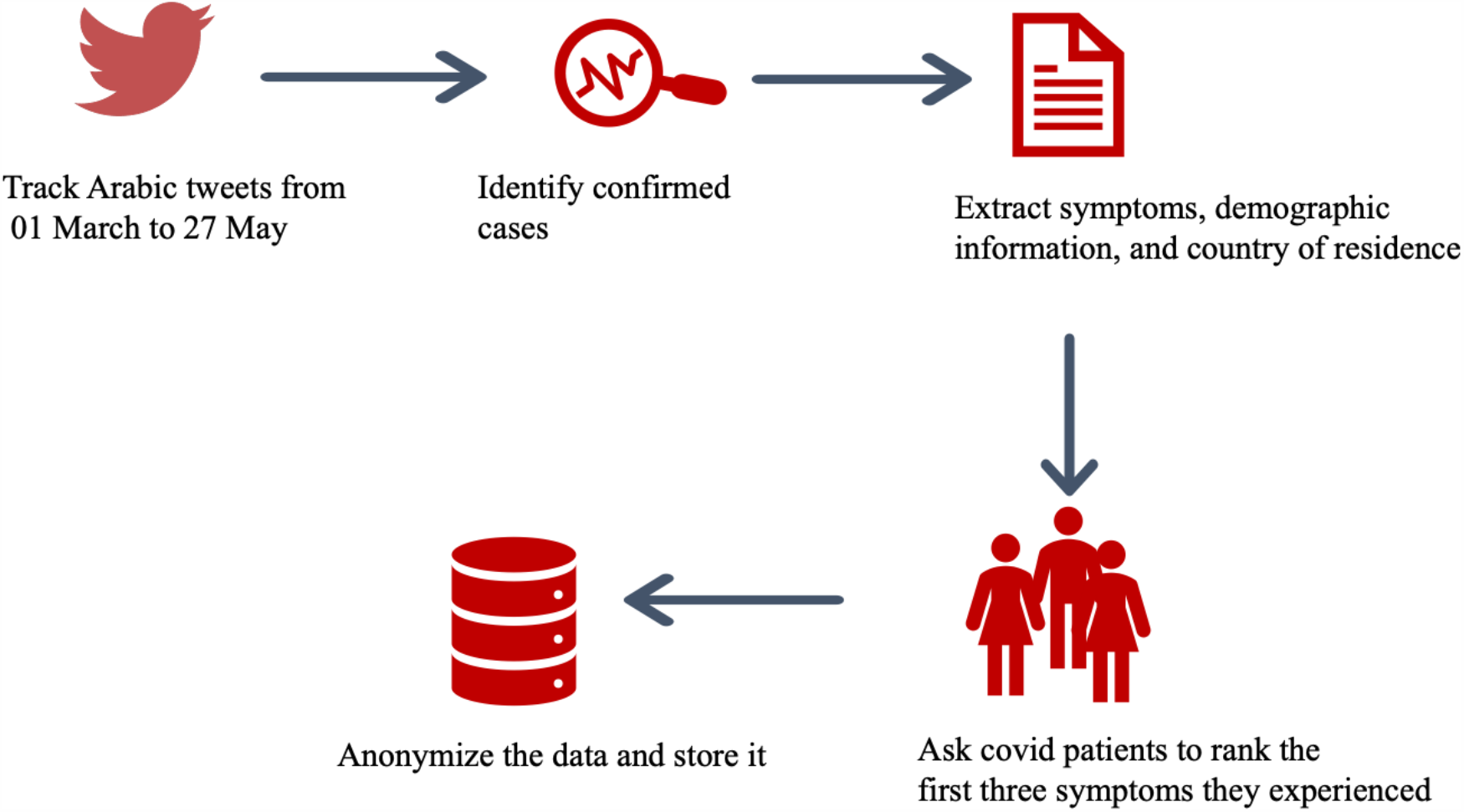
Data collection steps.

In this paper, we study COVID-19 symptoms as reported by Arabic tweeters. Initially, we shuffled Arabic tweets and searching for tweets with COVID-19 symptoms and also collected tweets for users who reported themselves infected through clinical test. In addition, we asked users who have been marked infected about the first three symptoms they experienced via a voluntary survey template sent over private message.

## Methods

Our method for data collection is outlined in Figure 1. We search Twitter for personal reports of covid-19 from March 1^st^, 2020 to May 27, 2020 using two Arabickeywords 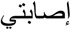 and 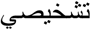 which translates roughly to *“I have been diagnosed”*. Suck keywords are likely to filter out reports that were not associated with a formal test result. An initial list of 463 users were collected and two independent freelancers were asked to further read users’ timeline and extract symptoms that were explicitly mentioned to be related to covid and their order of appearance if mentioned. Additional information such as user gender, date of infection, and the country of residence were also collected. We assume the date of tweeting being covid-19 positive is the date of infection in case no other information were available.

A final list of 270 covid-19 users were identified amongst 80 were sharing their symptoms publicly. To further understand the chronological order of the symptoms, we asked through Twitter personal messages users to order the first three symptoms they experienced right before or after tested positive for covid-19.

We record the symptoms from first to last based on the received responses and what is available publicly on the users’ tweets. In case no order was given, an implicit order is assumed following the order of which the symptoms were mentioned by the user.

Tracking tweets containing specific keywords is simply not enough to have the big picture about the disease dynamics [9]. Many patients detail their experience while infected, hence, knowing their health condition, sentiment, and tracking useful information may lead to a better understanding of the disease symptoms. In particular, we found tweets that were posted within ±5 days of infection date to contain valuable information about early symptoms, allowing us to process and rank the symptoms. Figure 3 highlights three tweets by three different covid-19 patients that indirectly relate symptoms before or after diagnosed with covid-19. For simplicity, we set a fake date (28 Apr, 2020) for all the three using the TweetGen service [10]. User..1 was tested positive on Apr 26 and tweeted on Apr 28 about the loss of smell. User…2 tested positive May 1^st^, three days after complaining about a headache. User..3 tested positive Apr 29^th^, one day after tweeting her wish to be able to taste the food.

**Figure 3:**
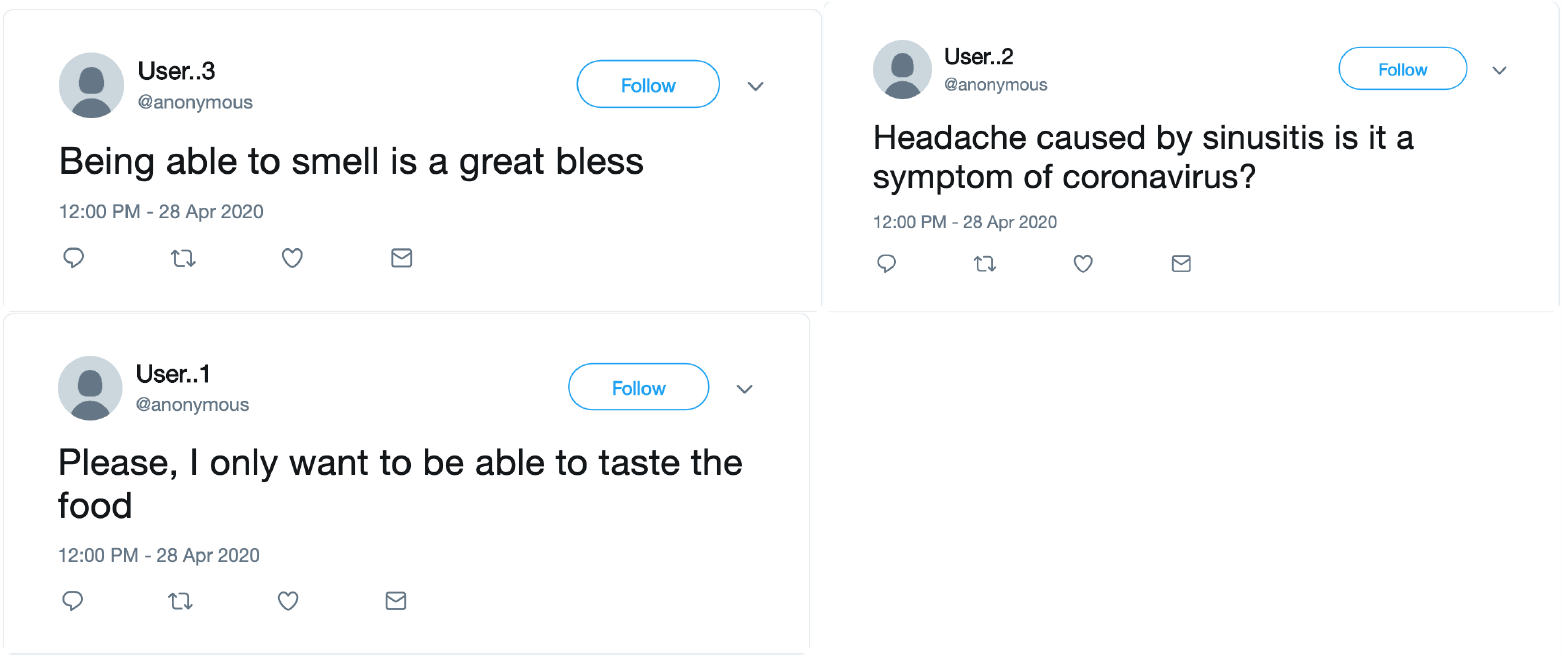
Example of tweets collected within ±5 days of the user tweeting being covid-19 positive

The example highlighted in Figure 3 demonstrates that mining twitter for covid symptoms require more than a simple keyword search. In principle, the context of the tweet, narrated by a covid-19 patient, is also important. Therefore, it is important to look not only the verbatim of the tweet but also to its context. To build a high-quality database of covid symptoms based on Arabic tweets, we have relied on manual symptoms extraction. However, we have used Twitter API to construct a social network graph for the 270 users and the software Gephi [11] to visualize the resulted graph.

## Results

The majority of the cases were recorded in May (78%, n=210), followed by April (14%, n=39) and March (8%, n=21). This surge of May reports is understandable as most of world countries, let alone the Arabic speaking countries, witnessed a great increase in the number of confirmed cases. As for the demography, users from Saudi Arabia, Kuwait, and UAE constitute 85% of reports with Saudi Arabia being the largest cluster of reports (46%). The other countries (Egypt, Iraq, Bahrain, Qatar, UK, USA, Belgum, and Germany) together constitute the remaining 15%. Needless to say, some of the adopted strategies to prevent further spread of the virus (e.g., active screening by the Ministry of Health in Saudi Arabia [12]) may also helped in finding more reports in May compared to other months. We have witnessed this firsthand as some of the asymptomatic reports were mainly a result of early active screening.

The fact that almost half of the reports came from Saudi Arabia is not surprising as its one of the top countries participating on Twitter with more than 15 million users [13]. We have collected almost 900 symptoms from the 270 reports (as shown in Figure 4). The daily number of collected tweets is also highlighted in Figure 5.

**Figure 4:**
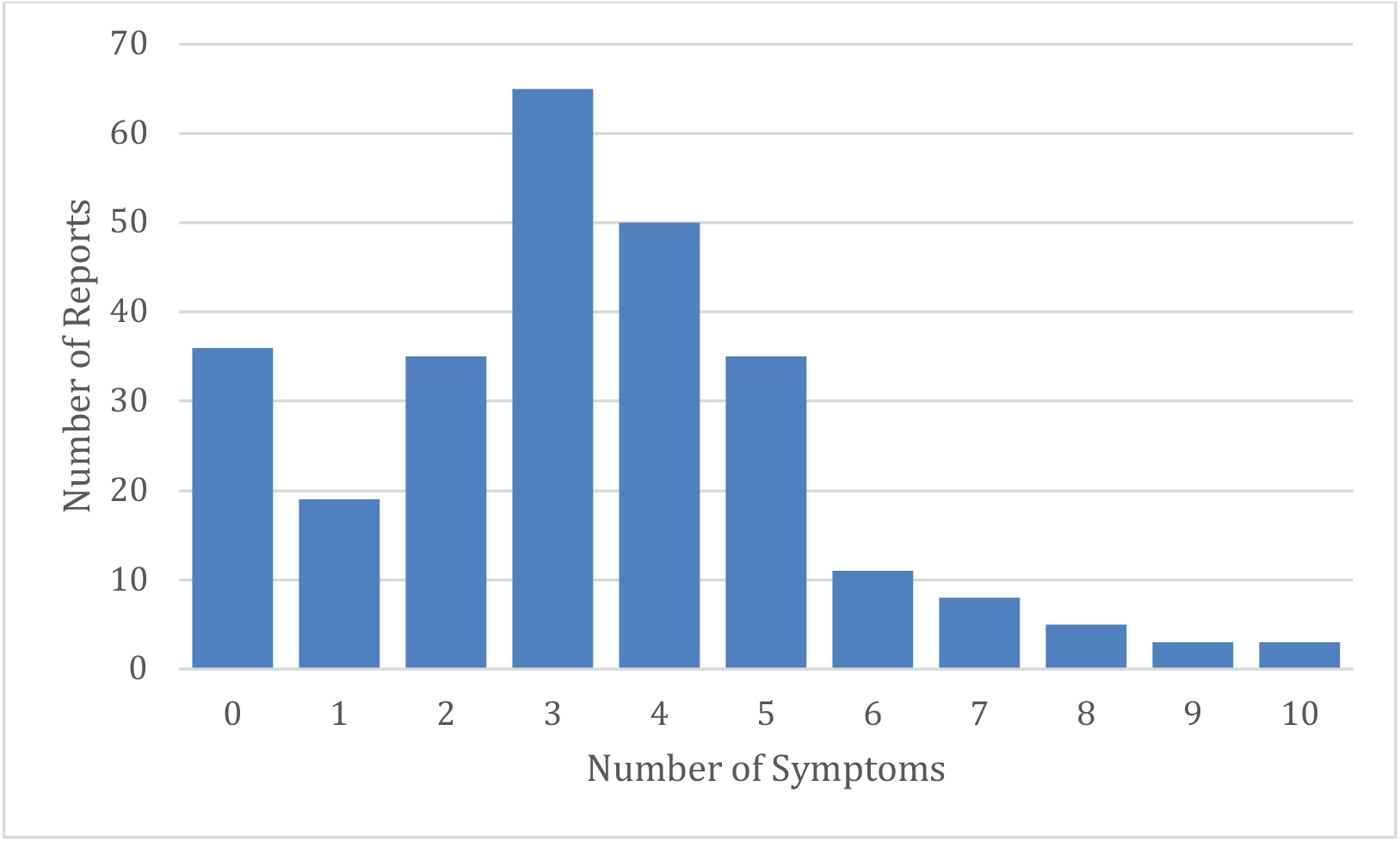
Number of reports distributed by number of symptoms.

**Figure 5:**
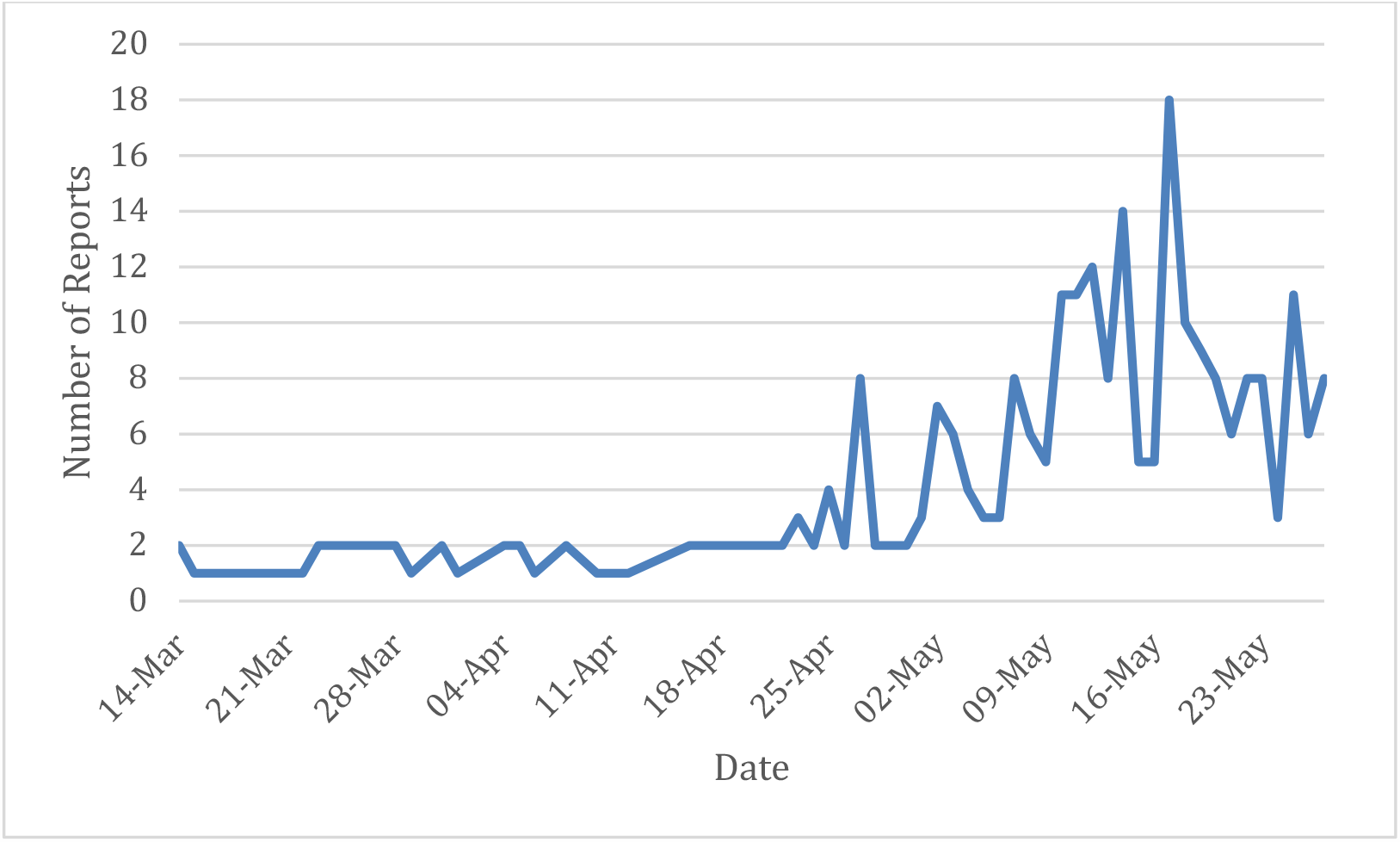
Number of daily collected reports from Twitter for March to May 2020

Figure 4 indicates that most of the reported cases experience between 2 to 5 symptoms, whereas 13% of the reported cases were asymptomatic. Table 1 lists the frequency of each symptom ordered from the most prevalent symptom to the least. Only Fever experienced by more than 50% of the patients. The frequency of symptoms appears to be consistent across male and female patients (Corr. Coeff. =0.966). Further, Table 2 lists the top three most reported symptoms. Fever and headache were commonly the first reported symptoms. The top 4 symptoms that coincide with fever were headache (23.7%), cough (14.4%) anosmia (13.7%), and ageusia (12.2%). Other symptoms have relatively lower frequency with fever. In addition, Table 3 lists the top-8 common symptoms for Saudi Arabia and Kuwait which correspond for about 81.2 % of the reports. The symptoms have a Corr. Coeff. of 0.835 between the two countries.

**Table 1:** Most common symptoms reported by the users

| Symptom | All users<br>n=234 (%) | Male<br>n=171(%) | Female<br>n=63 (%) |
| --- | --- | --- | --- |
| Fever | 139 (59) | 98 (57) | 41(65) |
| Headache | 101 (43) | 68 (40) | 33(52) |
| Anosmia | 91 (39) | 63 (37) | 28(44) |
| Ageusia | 72 (31) | 51 (30) | 21(33) |
| Fatigue | 68 (29) | 54 (32) | 14(22) |
| Cough | 62 (26) | 48 (28) | 14(22) |
| Sore Throat | 42(18) | 30 (18) | 12(19) |
| Dyspnea | 33(14) | 26 (15) | 7(11) |
| Diarrhea | 27(12) | 22(13) | 5(8) |
| Runny Nose | 23(10) | 17 (10) | 6(9) |
| Arthralgia | 16(7) | 10 (6) | 6(9) |
| Chest Pain | 15(6) | 13(8) | 2(3) |
| Back Pain | 14(6) | 11(6) | 3(5) |
| Anorexia | 14(6) | 11(6) | 3(5) |
| Body Ache | 12(5) | 8(5) | 4(6) |
| Nausea | 12(5) | 8(5) | 4(6) |
| Osteodynia | 11(5) | 8(5) | 3(5) |
| Dry throat | 9(4) | 6(3) | 3(5) |
| Myalgia | 9(4) | 7(4) | 2(3) |
| Dizziness | 8(3) | 6(3) | 2(3) |
| Chills | 7(3) | 5(3) | 2(3) |
| Nasal Congestion | 7(3) | 4(2) | 1(2) |
| Sinusitis | 7(3) | 3(2) | 4(6) |

**Table 2:**
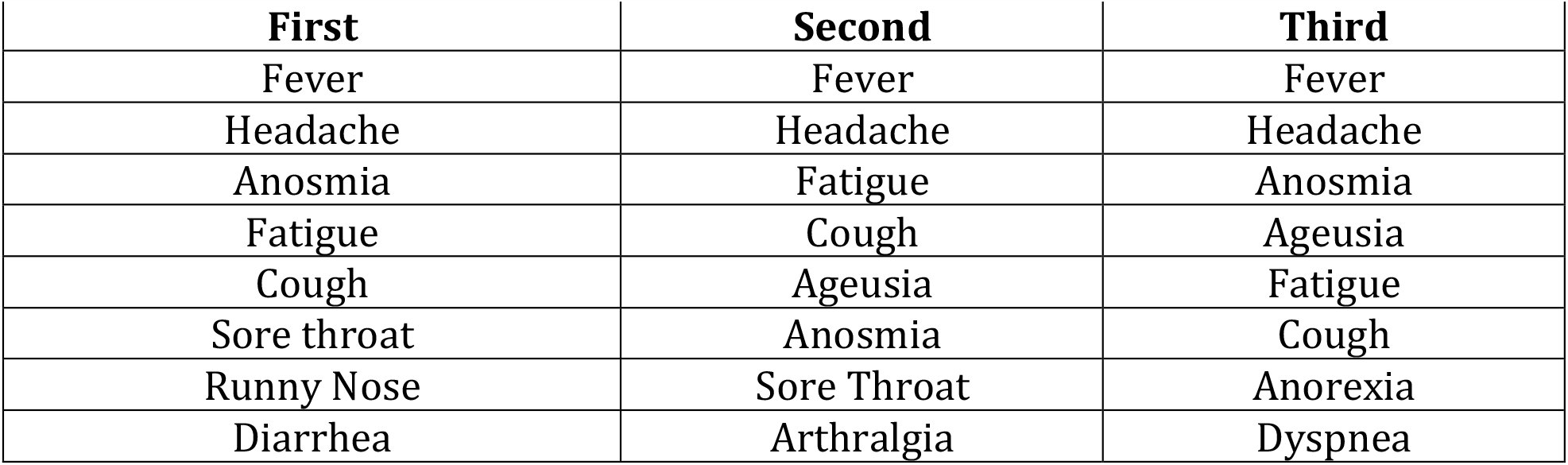
Top-8 first, second, and third symptoms reported by the users

**Table 3:** Top-8 common symptoms for Saudi Arabia and Kuwait

| **Symptom** | **Saudi, n=110 (%)** | **Kuwait, n=80 (%)** |
| --- | --- | --- |
| Fever | 65 (59) | 45 (56) |
| Headache | 42 (38) | 38 (48) |
| Anosmia | 46 (42) | 21 (26) |
| Ageusia | 36 (37) | 19 (24) |
| Fatigue | 31 (28) | 19 (24) |
| Cough | 21 (19) | 19 (24) |
| Sore Throat | 22 (20) | 11 (14) |
| Dyspnea | 14(13) | 11 (14) |

We have constructed a social graph for the users to discover insights related covid-19 communities in social networks as shown in Figure 6. An edge between X and Y represents the fact that X follows Y or Y follows X or both follow each other on Twitter. Saudi cases were colored in green while the Kuwait cases are blue and orange for other places of residence. The graph shows 164 nodes each node with a size proportional to its degree in the graph. The remaining users were found to be disconnected from all other users and thus removed from the network. It was built by the software Gephi [11] and relations were extracted by Twitter API. Clearly, Kuwait users are more connected compared to others and some form a clique of three or more users.

**Figure 6:**
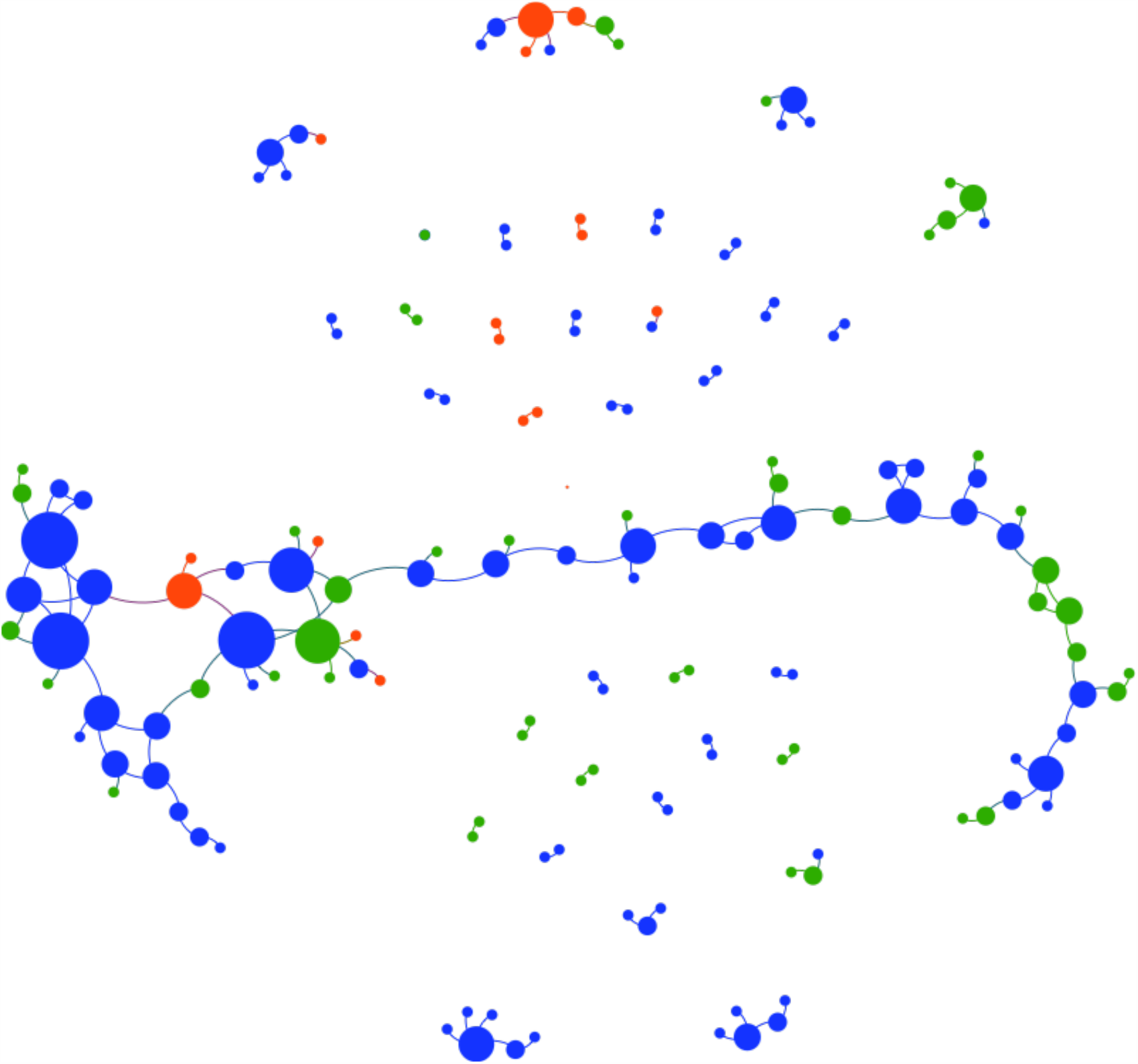
The social network graph of the users

## Discussion

This work identified common covid-19 symptoms from Arabic personal reports on Twitter. Such study complements other recent studies [5-6,9] that were focused on English tweets or specific demographic groups. The study was carried in a way to report not only the symptoms but their timeline as narrated by users. Social networks have become the de facto communication channel for large number of people. Many individuals around the globe write, interact, or even just browse the content of the social networks countless times a day. Social networks have the property of being continuously updated by new information provided by other global citizens. As such, it is crucial to monitor their content to identify health issues [14-15]. One potential benefit of analyzing social networks is understanding covid-19 symptoms and identifying people at high risk [7].

Anosmia being one of the top three reported symptoms, mentioned in 39% of the reports, was a surprising result in our study. Several tweets complained how long it lasted before they started to smell again. Our sample size is still relatively small to make any good judgement in this regard. However, recent clinical studies have reported finding Anosmia in 35.7% of the mild cases of covid-19 [16] which is relatively close to our estimation from Arabic tweets. Indeed, the size of self-reports reflect the testing capacity in different countries. As of June 9, 2020, Saudi Arabia has done almost one million tests and Kuwait has roughly exceeded 350,000 tests [17].

## Limitations

The reported cases from Egypt, the Arabic country with largest population, were inadequately representative. This could be attributed to factors such as the preferred social platform (e.g., Facebook), the dialect and use of local idioms.

Our study tracked two widely used keywords to identify Arabic covid-19 patients on Twitter then manually extract symptoms. More complex keywords could reveal more interesting patterns about symptoms, and this brings the need to establish a comprehensive medical dictionary for different local Arabic dialect. The dictionary can be utilized when mining different health opinions and conditions from the Arabic content in social networks.

Our main motivation is to extract symptoms from users who are likely took the disease test and, hence, tweeted based on its result. In this study, we have not used other covid-19 sources. Specifically, studying the Arabic content of personal reports from both Facebook and Twitter will enrich the study.

The noticeable increase in May reports compared to other months show the importance of developing a real-time surveillance system based on the symptoms reported by the Arabic content of Twitter. It also suggests further studies into the information sharing behavior [19] in different communities and.

## Conclusion

The collected tweets of symptoms provided some insights into covid-19 symptoms and their chronological order. We have shown the most common symptoms associated with Arabic self-reports of symptoms. We have analyzed the first, second, and third common symptom experienced by the users. Furthermore, we analyzed symptoms prevalence in the two largest clusters found in our database: Saudi Arabia and Kuwait.

## Data Availability

Data is continously updated and available upon request

## Acknowledgements

This work was supported by King Abdulaziz City for Science and Technology (Grant Number: 5-20-01-007-0033).

## Authors’ Contributions

Eisa Alanazi and Abdulaziz Alashaikh designed the study and wrote the manuscript. Sarah Alqurashi developed the social network analysis part and collected related tweets from the Twitter API. Aued Alanazi extracted and translated the symptoms from the collected personal reports to their scientific names. All authors approved the final version of the manuscript.

## Abbreviations

API: application programming interface
COVID-19: coronavirus disease

